# Machine-learning operations streamlined clinical workflows of DNA methylation-based CNS tumor classification

**DOI:** 10.1101/2024.01.25.24301176

**Authors:** Alexander L Markowitz, Dejerianne G Ostrow, Chern-Yu Yen, Xiaowu Gai, Jennifer A Cotter, Jianling Ji

## Abstract

**Background:** The diagnosis and grading of central nervous system (CNS) tumors, which was traditionally relied on histology, has been enhanced significantly by molecular testing, including DNA methylation profiling, which has been widely adopted for tumor classification. Clinical laboratories, however, are hindered when changes, such as the introduction of the Illumina Infinium MethylationEPIC v2.0 BeadChip, make existing classifiers incompatible due to shifts in targetable CpG sites among array versions. The aim of this study is to provide a scalable CNS tumor classification solution that empowers molecular laboratories and pathology teams to respond swiftly to these challenges.

**Methods:** We employed machine-learning operational methods including continuous integration and continuous training using 228 in-house MethylationEPICv1 array samples and two publicly available data sources to train and validate a DNA-methylation CNS classification pipeline that is compatible with Methylation450k, MethylationEPICv1, and MethylationEPICv2 BeadChips. We optimized CNS tumor classification by validating a multi-modal machine-learning classifier using a combination of a random forest and k-nearest neighbor model framework.

**Results:** We demonstrated an increase of accuracy, sensitivity, and specificity of CNS classification at the superfamily, family, and class level (class-level AUC = 0.90) after employing machine-learning operational methods to our clinical workflow. Our classification pipeline outperformed the DKFZv12.8 classifier in classifying pediatric CNS tumor types and subtypes when using the Illumina Infinium MethylationEPIC v2.0 BeadChip (concordance = 92%).

**Conclusion:** By leveraging machine-learning operational principles, we demonstrate a practical clinical solution for clinical molecular laboratories to employ for improved accuracy and adaptability in DNA methylation-based CNS tumor diagnostics.

**Importance of the Study:** Clinical molecular laboratories, neuro-oncology, and pathology diagnostic teams that utilize machine-learning classification systems are challenged when changes in underlying molecular technology make current systems inoperable. Our study provides a solution to clinically validate DNA-methylation profiling of central nervous system (CNS) tumors by employing machine learning operations, with a solution that is applicable to data generated from MethylationEPIC v2.0 BeadChip, as well as earlier versions. We show how continuous integration of novel data sources and algorithmic optimization substantially improves the robustness of the diagnostic tool and enables clinical laboratories to be agile in the face of evolving technology. Furthermore, we provide the computational infrastructure to scale out these services to any diagnostic laboratories focused on supporting CNS tumor classification.

**Key Points:**

- Our study addresses the crucial need for agility in clinical diagnostics, presenting a machine learning-enhanced CNS tumor classification pipeline that swiftly adapts to new technologies like the MethylationEPIC v2.0 BeadChip, ensuring seamless integration into existing clinical workflows.
- Utilizing machine learning operational methods, we curated an expansive reference dataset, leading to the optimization of our classification algorithm, which demonstrates superior adaptability and precision in CNS tumor diagnostics, essential for timely and accurate patient care.

## Introduction

Central nervous system (CNS) tumor diagnostics has a traditional reliance on histology but has evolved to encompass more contemporary strategies in molecular testing, which may reveal DNA sequence variants, gene fusions, copy number alterations, as well as distinct DNA methylation and RNA expression patterns (Louis et al., 2021; Hiemenz et al., 2018; Thomas et al., 2016). Among these, DNA methylation patterns, specifically the modification of cytosines in CpG sites throughout the genome, has emerged as a valuable diagnostic tool for tumor classification (Gielen et al., 2022). Distinct DNA methylation patterns, often specific to individual tumor type, have been described in recent years, leading to the widespread adoption of DNA methylation profiling and its acceptance as an invaluable molecular pathology tool, particularly in the classification of CNS tumors (Capper et al. 2018; Wu et al. 2022).

The widespread use of methylation profiling in tumor classification has been primarily benefited from the efforts of the German Cancer Consortium (DKTK) and German Cancer Research Center (DKFZ), who demonstrated the diagnostic accuracy of a machine learning classifier of CNS tumors and released a dataset of over 2,800 samples collected using the Illumina methylation array technology and demonstrated the diagnostic accuracy of a machine learning classifier for CNS tumors (Maros et al., 2020; Capper et al. 2018). Notably however, with the recent introduction of the Illumina Infinium MethylationEPIC v2.0 BeadChip (Noguera-Castells et al., 2023, Kaur et al., 2023), there is a need to update this classification system to account for the differences in targetable CpG probes following the technological transition. This presents a real-world challenge for machine learning classifiers including this CNS tumor clinical utility, as they are at risk of becoming inoperable when significant changes are made to the set of available CpG probes. Previous studies that have investigated methods to validate a methylation-based classifier in the context of the 2021 WHO Classification of CNS tumors did not employ methods to accommodate inevitable shifts in data structure and availability (Santana-Santos et al. 2022; Chen et al., 2021; Maros et al., 2020). While updated model metrics from the latest DKFZ classifier are not publicly available, many clinical laboratories that validated their classification pipelines end up using dated results to evaluate their own models. Nonetheless, as we would like to show in this study, access to the web-based DKFZv11-12 CNS classifiers and alternate publicly available datasets provide the necessary tools to address this clinical problem if approaching it differently as we describe below.

Here, we provide a scalable CNS tumor classification system compatible with Illumina Infinium HumanMethylation450 BeadChip (450K), Infinium HumanMethylationEPIC (EPICv1), and Infinium HumanMethylationEPIC version 2.0 (EPICv2) arrays. By applying machine-learning operation (MLOps) principles, we demonstrate how this system can be continuously augmented and validated in response to inevitable changes such as technological and algorithmic advancements. We assessed the DNA methylation profiles of a hold-out test set of EPICv2 array samples to show how improvements to the CNS classification system can be achieved with continuous integration of data sources and algorithmic optimization. We view this as an adequate response to the need of decentralizing machine-learning operations of this clinical utility and promote a more transparent assessment of methylation-profiling of CNS tumors. In addition, our comprehensive study shows how the current standards can be further improved by using a hierarchical machine-learning classification system.

## Methods and Materials

### Data Collection

We aggregated DNA methylation array datasets of CNS tumor samples from a combination of public and in-house sources including the Gene Expression Omnibus (GEO) database. Our study procured DNA methylation data generated from the Illumina Infinium HumanMethylation450K BeadChip (450K), Infinium HumanMethylationEPIC (EPICv1), and Infinium HumanMethylationEPIC version 2.0 (EPICv2) arrays (Illumina, USA), technology designed to capture the epigenomic landscape of previously tested CNS tumor samples (Capper et al., 2018). We separated our data into a training, validation, and testing dataset. The training dataset consisted of 3029 raw data files (.idat) derived from GSE90496 (n = 2801 samples), GSE141039 (n = 153), and 228 in-house (CHLA) samples. The validation set (GSE109379) consists of 1104 samples. The hold-out testing set consists of IDAT data of 42 samples, collected with the EPICv2 array, including limit of detection (DIL) and repeatability runs (WD, IOP) (**Supplemental Table 5**). All data was collected in compliance of policies set by our institutional review board.

### Data Preprocessing

DNA was extracted from frozen tumor tissue using the Gentra Puregene Tissue Kit, and from FFPE using the QIAamp DNA FFPE Tissue Kit. All DNA was RNAse treated and quantified with the Promega Quantus. Quality of DNA from FFPE samples was assessed via qPCR using the Infinium HD FFPE QC Assay. The Zymo EZ DNA Methylation-Lightning Kit was used to bisulfite convert and purify DNA. For FFPE-derived samples, the Illumina Infinium HD FFPE Restore protocol was used to repair any degradation prior to proceeding with the Infinium assay. Following the restoration, all samples were amplified, fragmented, precipitated, and resuspended using the Infinium Methylation EPIC kit. Samples were hybridized to Infinium MethylationEPIC BeadChips and single-base extension was performed followed by staining. Arrays were imaged using the iScan instrument to generate IDAT files for downstream analysis.

Bioinformatic processing of IDAT files were conducted in the R programming environment (v4.2.1). We used minfi (v1.42.0) (Aryee et al., 2014) library to import and process IDAT files. Standard background correction and dye corrections were performed by parameter setting the average probe intensity to 10,000 for red and green channels. We performed batch correction for formalin-fixed paraffin-embedded (FFPE) and frozen samples (KYRO/Frozen) using the limma (v3.52.4) library (Ritchie et al. 2015). We corrected for batch effects of the validation and test sets using the calculated batch effect of the training dataset. We calculated beta-values, as recommended by Illumina, by dividing adjusted methylated values by the sum of the adjusted methylated and unmethylation values with an offset of 100.

### Machine-learning classification of CNS tumors via DNA methylation profiling

In our pursuit of achieving the most granular and informative categorization of CNS tumor samples, the MMAA pipelines utilize a hierarchical classification system that mirrors the 2021 WHO CNS tumor classification hierarchy (Louis et al., 2021). A key challenge in this endeavor was the unavailability of a publicly accessible dataset that was annotated and aligned with the 2021 WHO classification hierarchy, refined based on methylation data. This prompted us to thoroughly annotate the reference dataset with WHO classifications extending across the superfamily, family, and class levels. (**Supplemental Table 1**, Capper et al., 2018).

### CpG probe selection for hierarchical and non-ordinal classification

For the hierarchical classification system, we wanted to refine the CpG probes used when predicting at each level. First, a set of CpG probes were selected based on the highest variance of beta-values among the entire reference dataset. We then iterated this process at the superfamily level, where we assessed the highest variance of beta-values per superfamily to select probes to predict linked families. Finally, we iterated the process again at the family-level to select CpG probes most varied of the linked classes. This process results in a classification pipeline that considers a total of 44 sets of 10,000 CpG probes, selected to target the hierarchical classes at the superfamily, family, and class levels of the 2021 WHO Classification of CNS tumors (**Supplemental Table 2**).

In addition to our hierarchical classification system, we recapitulated a random forest model using the methods described by Capper and colleagues previously (Capper et al., 2018). The random forest model used a set of 10,000 CpG probes based on the importance coefficients of a random forest varsel of the training dataset exclusively derived from the GSE90496 cohort. We found 1,240 CpG probes that were not applicable with the EPICv2 array. We explored the use of a random forest model (RF8760) using the remaining 8,760 CpG probes that were universally compatible with all array types. The RF8760 model was trained using the beta-values processed using a non-ordinal classification model, with a fixed set of CpG probes for all cancer types.

### A series of machine-learning classifiers are trained to provide hierarchical classification

We trained of a suite of supervised-learning models, corresponding to each hierarchical level. Our Multi-modal Methylation Array Analysis (MMAA) pipeline is comprised of both a random forest (RF) and a k-nearest neighbor (KNN) model, which were used in conjugation in our machine-learning pipeline to generate predictions. CNS tumor classification predictions are determined by the highest predictive score, whether it emanated from the RF or KNN model. To refine and enhance the predictive accuracy of our models we calibrated the raw scores using a cross-validated generalized linear model, a process that was fine-tuned with default regularization parameters and described by Capper and colleagues (Capper et al., 2018). This calibration stage generated the final classification and score for each CNS tumor sample under consideration.

When predicting the CNS tumor classification, the pipeline leverages a total of 26 sets of RF and KNN models, each responsible for generating predictions at a distinct nodes within the hierarchical classifications. To begin, a superfamily is determined by utilizing a set of the top 10,000 variable CpG probes. Once the superfamily is identified, the prediction process continues by selecting a CNS family, which is based on a set of 10,000 CpG probes that exhibit the highest variability within the chosen superfamily. Subsequently, the process is reiterated at the class level, predicting the class using a new set of 10,000 CpG probes with the highest variability at the class level, specifically tailored to the predicted family. In cases where a superfamily or family contains only one class, the predicted class is determined based on the level immediately above it in the classification hierarchy.

### Interpretation of classification scores

All scores generated by the MMAA pipeline consist of a calibrated classification score, which reflects the prediction probability of being assigned a classification. The superfamily score gauges the efficacy of classifiers in predicting superfamily classification, with 1 being the highest attainable score. Similarly, the family and class scores measure the classifier’s accuracy in predicting family- and class-level classification. Notably, a score above a threshold of 0.9 suggest a match in classification. A score between 0.6 through 0.9 would be interpreted as a suggestive classification. Scores below 0.6, would be inconclusive and would be interpreted as non-applicable classification.

### Continuous training, integration, and deployment of methylation-based CNS classifier

Our computational pipeline employs machine learning operational principles of continuous integration/continuous deployment (CI/CD) to provide robust and reliable classification services. CI/CD integration involves automating the pipeline from data acquisition to model training and deployment (**Figure 1**). By implementing CI/CD protocols, clinical laboratories can ensure that their classifiers remain up to date with the latest data, methodologies, and clinical insights, which translates to enhanced accuracy of their diagnostic workflows.

**Figure 1.**
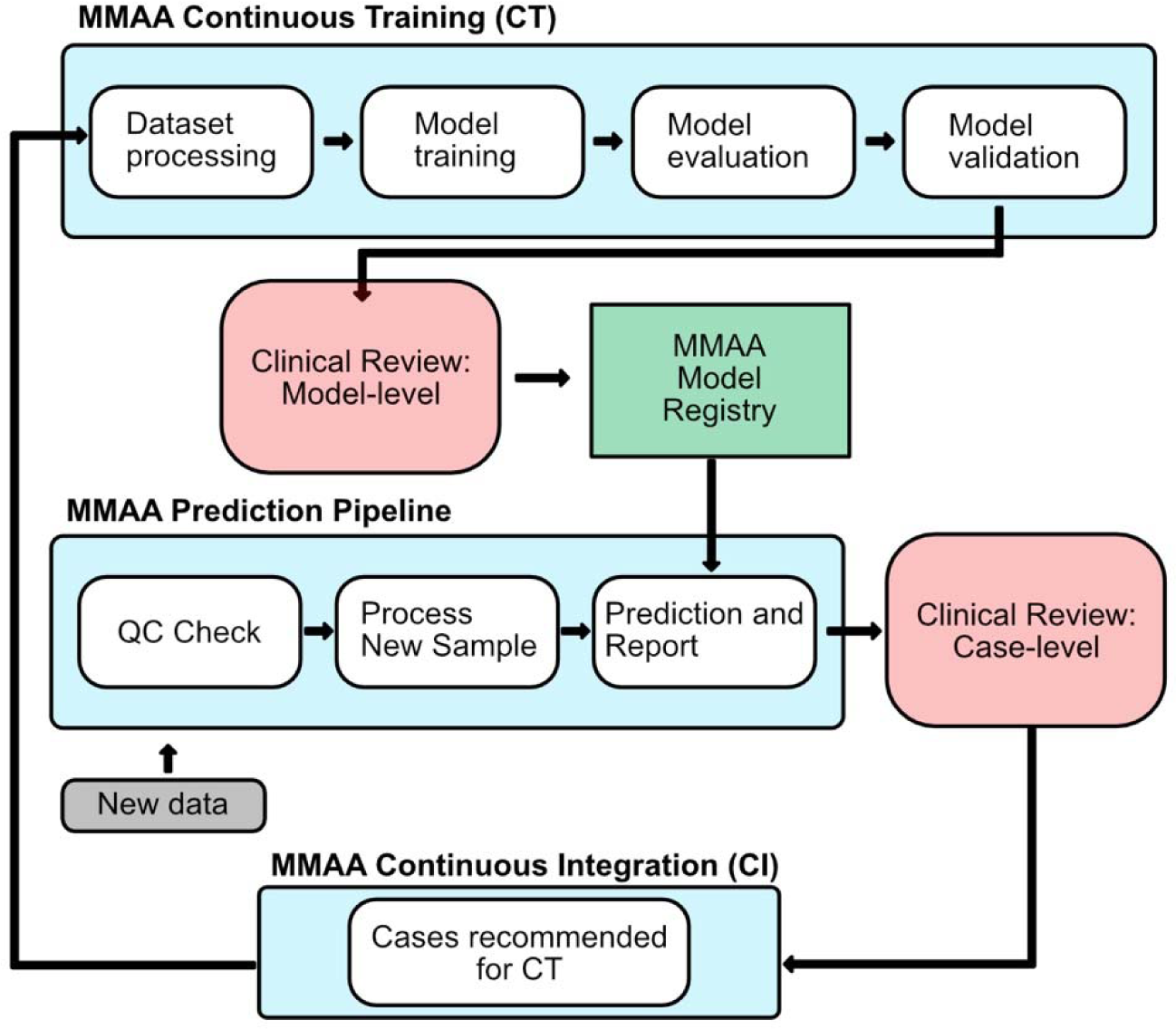
Clinical machine-learning operations for DNA methylation based central nervous system (CNS) tumor classification. Public and in-house cases are aggregated into a reference dataset for model training. A clinical review to evaluate model performance includes assessment of sensitivity, specificity, and accuracy of classifications of a validation dataset and an in-house testing dataset. An accepted model is registered for case CNS predictions services. Model monitoring at the case-level is performed to recommend candidate cases for continuous integration of the reference dataset. Employing machine-learning operations during the clinical review at the model-level and case-level enables rapid assessment of the CNS classification system.

Continuous integration happens at the predictive and training levels of our clinical workflow. At the predictive level, new methylation data is assessed using our bioinformatics processes and machine-learning models. Having continuous integration allows for rapid case warehousing, reporting, identification of edge cases, and evaluation of model performance over time. Simultaneously and at the training level, continuous integration automates the retraining of our machine-learning models, triggered by expert consensus of the new data (**Figure 1**). This process enhances our clinical diagnostic pipeline to be agile in the face of novel methylation-profiles associated with integrated diagnoses, limitations in true negatives represented in the reference dataset, and changes to clinical nomenclature of CNS tumors.

Continuous training consists of training a series of hierarchical machine-learning models involves the use of validation and hold-out testing sets to evaluate model performance and drift. Expert consensus would require the validation of model metrics on independent hold-out test datasets including the original validation dataset provided by DKFZ (GSE109379, n=1104) and a curated in-house set with integrated neuro-oncology team consensus. We integrate new cases into the reference dataset (n=228) after a clinical review, selecting high-confidence cases based on an integrated pathology diagnosis. In-house cases that had a classification score above 0.9 were selected to be integrated into the reference dataset for continuous training. We assess model performance based upon the area-under-curve (AUC) metrics of a validation set of 1104 samples (GSE109379) and by assessing the accuracy on a hold-out test set of 42 samples. During the clinical validation, the model is only accepted for production after prior-determined sensitivity, specificity and accuracy. The criteria for model acceptance are an AUC metric >= 90% for the 1104 sample validation dataset and >= 90% concordance of machine-learning classification and integrated pathology diagnosis. These were chosen based upon what was commonly acceptable by the field at the time of the study (Santana-Santos et al., 2022).

### Statistical analysis and data visualization

All data was analyzed in the R programming environment (v4.2.1). Data visualization was performed using the BPG package (v6.1.0). DKFZ (v12.8) CNS classifications were determined by using the web-based application.

### Data and Code Availability

DNA methylation data is available through the Gene Expression Omnibus (GEO) database. Random forest models were trained using the randomForest (v4.7-1.1) R package. K-nearest neighbor models were trained using caret (v6.0) R package. Calibrated score models were trained using the glmnet (v4.1) R package. All code and model data has been deployed in a publicly at (https://github.com/alex-markowitz/pipeline-mmaa) and is available as an open-source pipeline via a ocker container (https://hub.docker.com/r/alexmarkowitz/mmaa_pipeline).

## Results

### Multi-modal Methylation Array Analysis (MMAA) pipeline shows optimal performance in classifying CNS tumors via DNA methylation array profiling with Illumina EPICv2 array

The presented machine-learning classification system, Multi-modal Methylation Array Analysis (MMAA), leverages a training dataset established by integrating DNA methylation array data from three sources including publicly available (GSE90496, GSE141039) data sources and in-house samples, totaling 3,029 samples. Comparing to the published DKFZ model, the reference data sets are significantly enhanced in representation of atypical teratoid rhabdoid tumors, with 153 samples integrated from GSE141039, that of embryonal and glioma superfamilies with the inclusion of 228 samples (**Figure 2, Supplemental Table 3**). The remaining training data was derived from GSE90496, which added 2,801 DNA methylation array samples from yet another 91 cancer types and subtypes.

**Figure 2.**
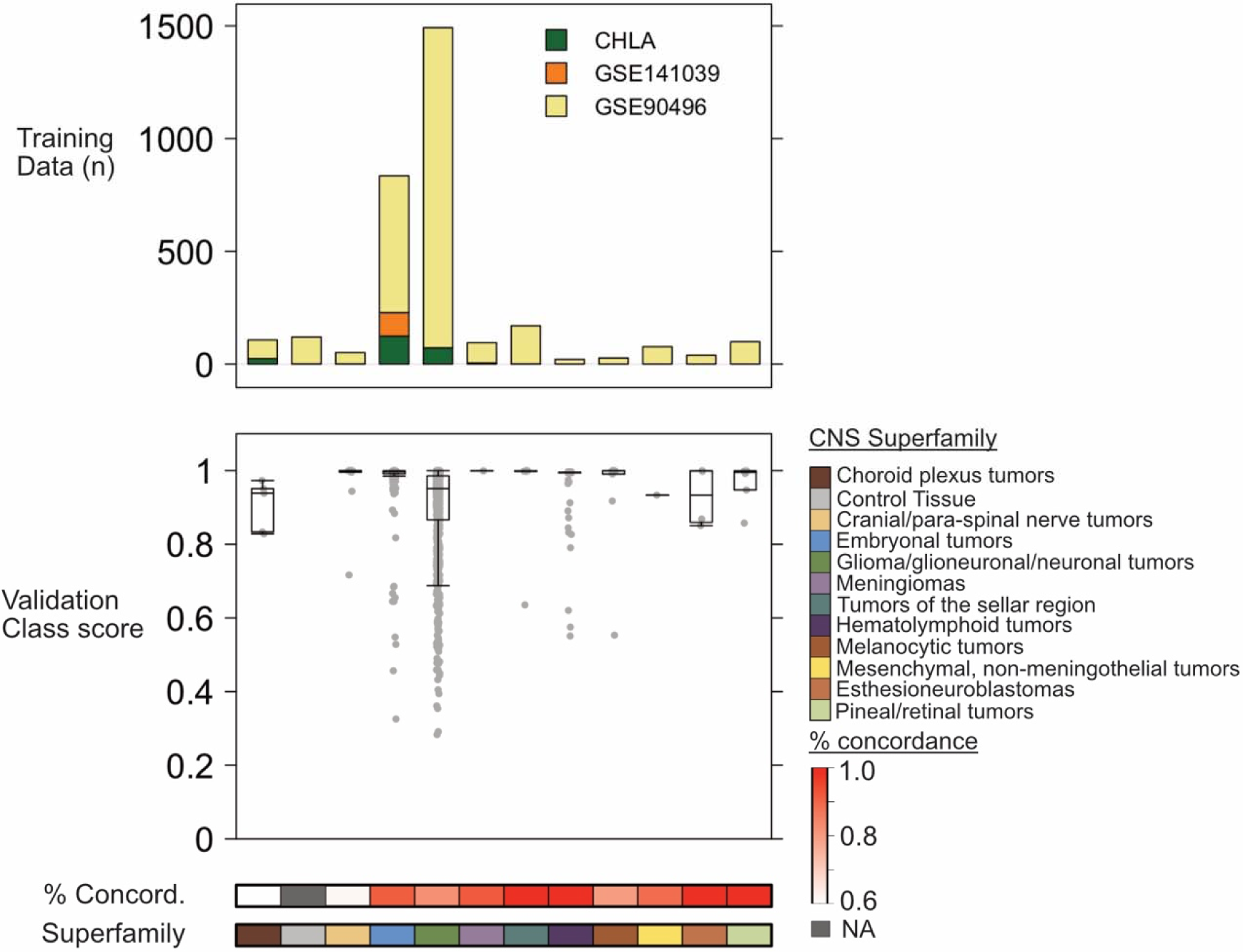
Distribution of CNS tumor samples among assigned superfamilies of the integrated reference dataset. The reference dataset consists of aggregating DNA methylation array data from publicly available sources (GSE90496, n = 2801; GSE141039, n = 153) and an in-house collection (CHLA, n = 228). Class prediction scores from a validation dataset (GSE109379, n = 1104) from the multi-modal methylation array analysis (MMAA) pipeline are displayed alongside the percent concordance between the EPICv2-incompatible random forest model (MAA) and the presented EPICv2-compatible MMAA pipeline. Choroid plexus tumor subtypes (0.6) and cranial/paraspinal tumor subtypes (0.6) displayed low concordance rates between models, suggesting that shifts in CpG probe availability among array types contributes to model drift.

The MMAA pipeline yielded an optimal performance in terms of specificity and sensitivity across the superfamily, family, and class levels of CNS tumor classification (**Figure 2-3**). The MMAA pipeline exhibited substantial discriminative power, as evidenced by area under the curve (AUC) values of 0.92, 0.94, and 0.90 for the superfamily, family, and class levels, respectively (**Figure 3**). The AUC values generated using the MMAA model closely mirrored those reported by Capper et al (Capper et al., 2018). Importantly, the MMAA model is compatible with Illumina Infinium Methylation450K, MethylationEPICv1, and MethylationEPICv2 Beadchip arrays.

**Figure 3.**
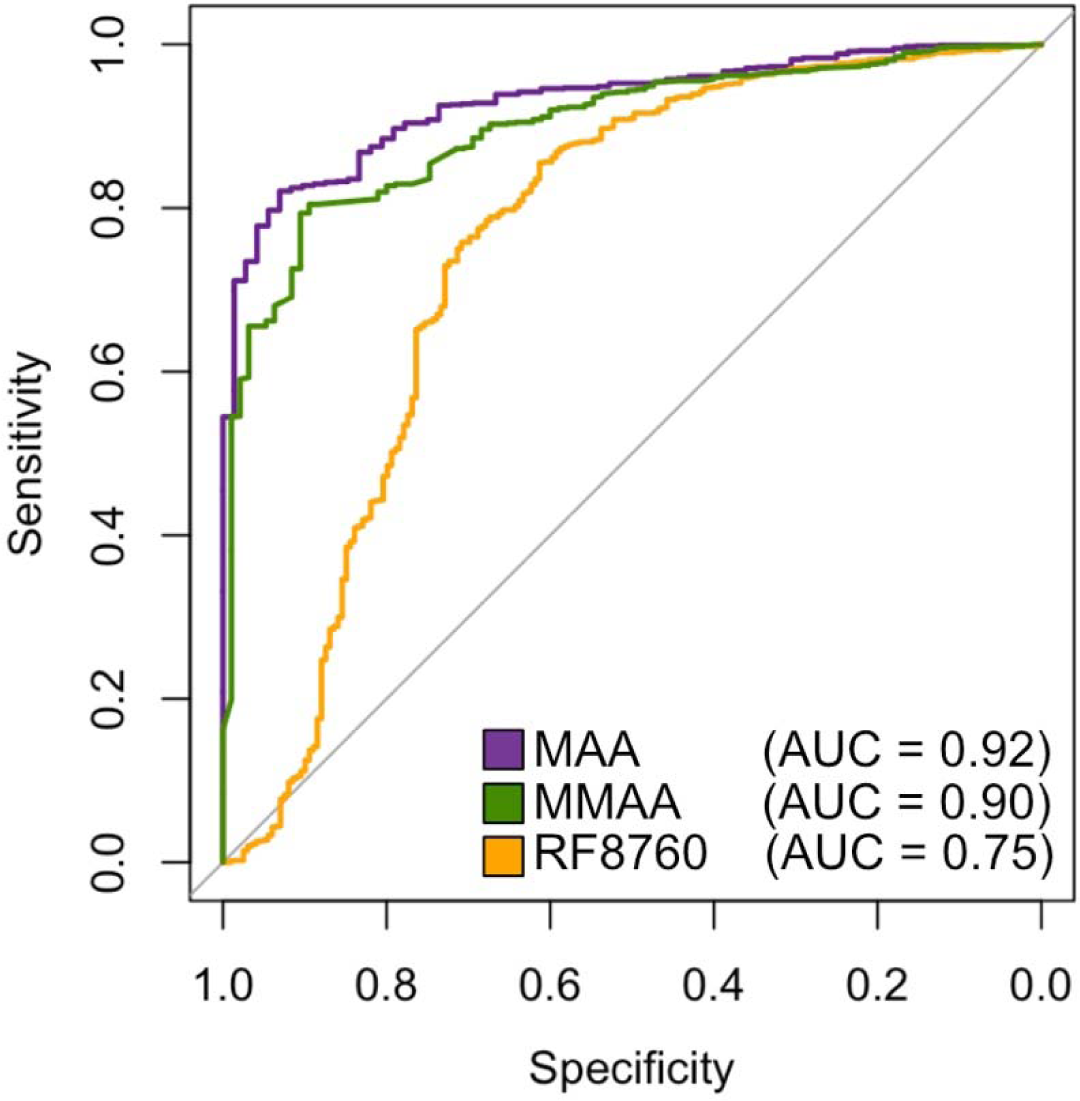
Receiver operational curves (ROC) comparing class-level model performance of three classification systems using the validation dataset (GSE109379, n = 1104). The three models compared include an EPICv2-incompatible random forest classifier (MAA, 10K) using 10,000 CpG probes (AUC = 0.92, purple), an 8,760 CpG probe-based model derived from the CpG probes of MAA that overlap with all array types (RF8760, AUC = 0.75, orange), and the Multi-modal Methylation Array Analysis (MMAA) pipeline (AUC = 0.90, green). The MMAA classification pipeline shows comparable results to the original classification pipeline and was recommended for clinical prediction services.

Comparing previous classification models is difficult due to the limitations of exclusively using CpG probes that overlap among all array types employed by these models. We investigated which machine-learning framework would be best for clinical production by evaluating metrics between three models: 1) a 10,000 CpG probe-based random forest model recapitulated the Capper et al methods (Capper et al., 2018) but not compatible with EPICv2 chips (MAA), 2) a 8,760 CpG probe-based model derived from the CpG probes of MAA that overlap with all array types (RF8760), and 3) the Multi-modal Methylation Array Analysis (MMAA) pipeline.

We report a slight decrease but comparable in model performance when comparing to a EPICv2-incompatible classification model (MAA) (**Figure 3**). We assessed class classifications concordance generated from MAA and MMAA of samples derived from the validation dataset (**Figure 2**). Concordance rates varied among superfamilies, with glioma, glioneuronal, and neuronal tumors showing the most varied scores (**Supplemental Table 4**). This shift in performance suggests that the CpG probes removed from the EPICv2 array were critical to classify a subset of samples in the validation dataset. Although, the CpG probes omitted from the EPICv2 array were chosen based on reliability, the majority of the reference data sets were collected using the Methylation450K array data, which is limiting for transferring selected CpG probes of interest to EPICv2-compatible models. Choroid plexus tumor subtypes (0.6) and cranial/paraspinal tumor subtypes (0.6) displayed low concordance rates between models. The imperfect concordance rate may suggest model drift, resulted from shifts in CpG probe availability among array types contributes to model drift. Low sample size of choroid plexus (n = 5), control tissue (n = 0), and spinal/para-spinal tumors (n = 13) may also contribute to observe concordance rates among superfamilies (**Supplemental Table 4**).

The MMAA pipeline significantly outperformed the use of a modified MAA random forest model (RF8760) that used 8,760 out of 10,000 CpG probes represented by all array types, where the AUC was notably lower at 0.75 (**Figure 3**). Here, we trained a random forest model using a subset of CpG probes from MAA, selecting the CpG probes that were compatible with 450K, EPICv1 and EPICv2 array types. This underscores the superiority and robustness of our approach, thereby affirming the reliability of our classifier for the accurate categorization of CNS tumor samples, especially in the context of the new EPICv2 array.

### Concordance rate of CNS classifications between MMAA and DKFZv12.8 CNS classification systems

We assessed the overall concordance of our MMAA pipeline with the DKFZ CNS tumor classifier (v12.8) using EPICv2 array chips (**Figure 4, Table 1, Supplemental Table 5**). The DKFZ classifier provided a non-applicable classification when the prediction score for a sample was below 0.3, as for 5 out of 42 (12%) samples generated. Of the remaining 37 viable samples, our MMAA classification pipeline produced 34 out of 37 (92%) family-level concordant classifications, with correlated prediction scores at the superfamily (Spearman’s ρ = 0.63, p = 2.7 x 10^-5^), family (Spearman’s ρ = 0.30, p = 7.45 x 10^-2^), and class (Spearman’s ρ = 0.71, p = 8.16 x 10^-7^) levels with the DKFZ v12.8 classifier.

**Figure 4.**
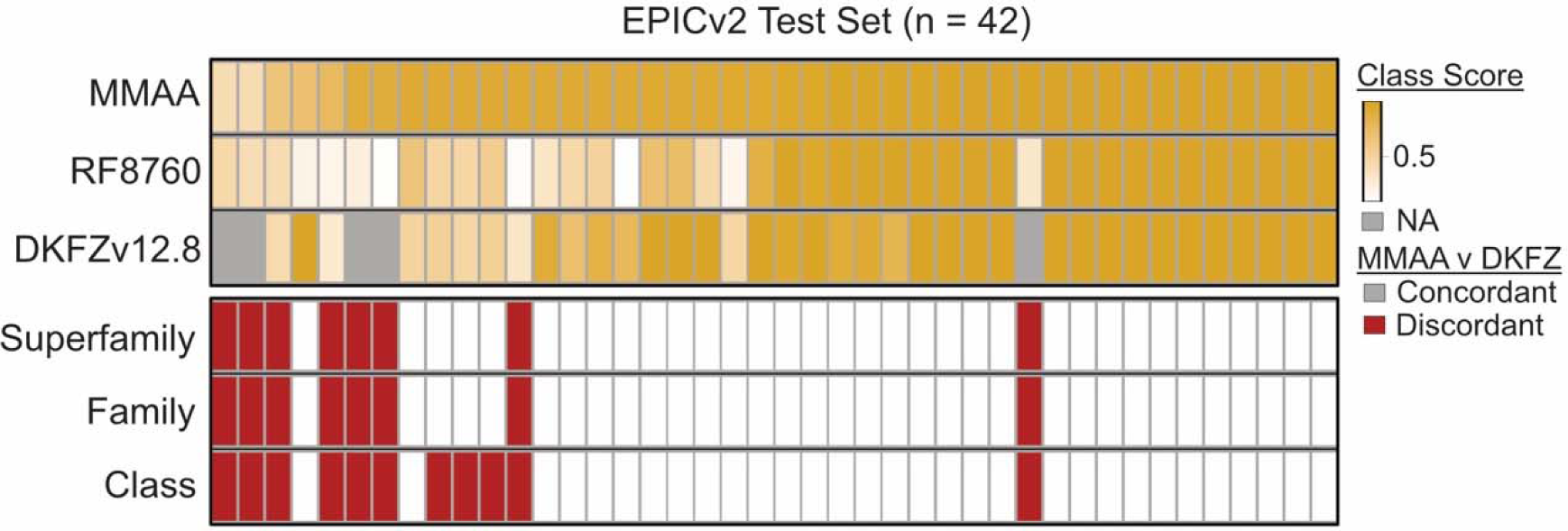
Class-level classification scores of a hold-out testing dataset of 42 Illumina EPIC v2 array chips using the MMAA pipeline, RF8760 model, and DKFZv12.8 CNS classification service. Class scores are arranged by the MMAA class score. DKFZv12.8 produced non-applicable values when a prediction score is below 0.3 (5 out of 42 samples). Class scores were significantly correlated among MMAA and DKFZv12.8 classifiers (Spearman’s rho = 0.71, p = 8.16 x 10^-7^). Discordant classification between MMAA and DKFZv12.8 were assessed at the superfamily, family, and class level. There were 34 (91.8%) cases where MMAA and DKFZv12.8 produced concordant family-level classifications.

**Table 1.** Discordant classifications between DKFZv12.8 and MMAA classification pipelines of samples using EPICv2 array. DKFZv12.8 does not generate an applicable classification if the prediction score is below 0.3 (5 out 11 samples). A neuroblastoma, *FOXR2*-activated sample (CHLA_16) was correctly classified via MMAA (0.99) but produced non-applicable results from DKFZv12.8. Control tissue samples showed discordances in 2 out of 4 samples. An ATRT sample (CHLA_36, CHLA_37, CHLA_ 38) ran three times for a reproducibility assessment and showed discordant class-level classifications between DKFZv12.8 and MMAA. There were 6 of 11 samples with scores below 0.9.

| CHLA ID | Sample Type | Integrated Pathology Diagnosis | MMAA Class | MMAA Score | DKFZv12.8 Class | DKFZv12.8 Score | RF8760 Score (EPICv2) | RF8760 Class (EPICv2) | MAA Class (EPICv1) | MAA Score (EPICv1) |
| --- | --- | --- | --- | --- | --- | --- | --- | --- | --- | --- |
| CHLA_16 | FFPE | CNS Neuroblastoma, FOXR2-activated | CNS NB, FOXR2 | 0.99 | NA | NA | 0.32 | GBM, RTK I | CNS NB, FOXR2 | 0.99 |
| CHLA_13 | FFPE | Cerebellum (non-tumor) | GG | 0.93 | MB, SHH | 0.33 | 0.12 | MB, SHH CHL AD | CONTR, CEBM | 0.79 |
| CHLA_36 | FFPE | Atypical teratoid/rhabdoid tumour | ATRT_TYR | 0.92 | ATRT_MYC | 0.52 | 0.48 | ATRT, MYC | ATRT, MYC | 0.95 |
| CHLA_37 | FFPE | Atypical teratoid/rhabdoid tumour | ATRT_TYR | 0.92 | ATRT_MYC | 0.47 | 0.47 | ATRT, SHH | ATRT, MYC | 0.95 |
| CHLA_38 | FFPE | Atypical teratoid/rhabdoid tumour | ATRT_TYR | 0.92 | ATRT_MYC | 0.53 | 0.56 | ATRT, MYC | ATRT, MYC | 0.95 |
| CHLA_6 | FFPE | Cerebral cortex (non-tumor) | CONTRL, HEMI | 0.89 | NA | NA | 0.24 | LGG, GG | CONTR, HEMI | 0.81 |
| CHLA_8 | FFPE | Cerebral cortex (non-tumor) | CONTRL, HEMI | 0.89 | NA | NA | 0.11 | LGG, GG | CONTR, HEMI | 0.86 |
| CHLA_25 | FFPE | High grade glioma (Lynch) | PA_CORT | 0.77 | GMB_RTK1 | 0.29 | 0.19 | GBM, RTK I | ANA PA | 0.27 |
| CHLA_24 | FFPE | Cerebral cortex (non-tumor) | DNET | 0.66 | CONTRL, HEMI | 0.43 | 0.41 | CONTR, HEMI | CONTR, HEMI | 0.82 |
| CHLA_21 | Frozen | High grade glioma | pedHGG_MYCN | 0.41 | NA | NA | 0.45 | GBM, RTK II | GBM,MYCN | 0.37 |
| CHLA_26 | FFPE | High grade glioma | pedHGG_MYCN | 0.41 | NA | NA | 0.40 | GBM, RTK II | GBM,MYCN | 0.37 |

There were 13 out of 37 cases where the DKFZ v12.8 classifier generated classifications with prediction scores less than 0.9. Of these 13 cases, 11 displayed discordant class-level classifications when compared to the output of the MMAA model. Notably, the DKFZv12.8 classifications matched the prediction of the RF8760 model which produced poor prediction results with only a 64% concordance rate (**Figure 4, Table 1, Supplemental Table 5**). Discordant predictions between MMAA and DKFZv12.8 did not directly reflect discrepancies with the integrated pathology diagnosis, as seen in 8 out of 11 samples of which MMAA generated an accurate classification (**Table 1**).

We assessed the cases where no classification could be generated using DKFZ v12.8 (EPICv2), yet our MMAA pipeline was able to generate predictions with prediction scores ranging from scores 0.41 – 0.99 (**Table 1**). Two non-tumor samples of non-tumor tissue sampled from the cerebral cortex were not correctly classified by the DKFZ v12.8 classifier; however, the MMAA pipeline correctly classified these as control tissue from the cerebral hemisphere (score 0.89, 0.89). In one case, a control cerebellum tissue was inaccurately classified as medulloblastoma by DKFZ v12.8 (score 0.33). Although the low-confidence score would not warrant clinical consideration, these discordant cases highlighted the need for the incorporation of diverse control samples in the reference dataset to account for the heterogeneity of methylation profiles of non-tumor samples.

In addition to inadequate classification of control samples among tumor samples, there were four additional samples that generated discordant calls using the EPICv2 array and DKFZ v12.8 classifier compared to our integrated pathology diagnosis and MMAA pipeline (**Table 1, Supplemental Table 5**). These included two samples of pediatric high-grade glioma, MYCN subtype (CHLA_21, CHLA_26). There was one high-grade glioma case (CHLA_25) with Lynch syndrome that generated low-confidence predictions from both MMAA and DKFZv12.8. Moreover, using the DKFZ v12.8 classifier with an EPICv2 array generated non-applicable predictions for a CNS neuroblastoma and FOXR2-activated sample that was previously validated via the EPICv1 and our MMAA pipeline (score 0.99). Similarly, a medulloblastoma and WNT-activated sample (CHLA_30) showed poor prediction power using the EPICv2 DKFZ v12.8 classifier (score 0.47), when previous classifications using a EPICv1 array-base classifier (MAA) and the MMAA pipeline with EPICv2 array confirmed this classification with high confidence (score 0.96). These cases highlight how MMAA improves accuracy of methylation-based CNS tumor classification.

## Discussion

The Illumina EPICv2 array, with updated content chosen to heighten the reliability and resolution of the assessment of methylation profiles, has had an unintended effect on diagnostic neuropathology, which has in recent years been increasingly relying upon methylation-based classification for clinical diagnosis. The launch of the EPICv2 array initiated a need for agility in clinical laboratory diagnostic testing, exemplified by a CNS tumor classification service that can withstand shifts in available CpG probe targets.

In response to this clinical operations problem, our assessment of the presented multi-modal methylation analysis (MMAA) pipeline with the EPICv2 array demonstrated its capability to yield highly accurate and reliable classifications, highlighting its potential as a valuable tool for the precise characterization of CNS tumor samples. Employing machine learning operations enabled us to streamline the integration of new data sources and optimize machine-learning algorithms. Furthermore, we showed how algorithmic experimentation may provide concordant predictions to use as a benchmark in implementing new machine-learning classifiers into the clinical setting. Employing both random forest and k-nearest neighbor models within an ordinal-classification framework is proven to increase performance in generating concordant classification of CNS tumor types and subtypes.

Validation of our improved DNA-methylation classification system maintained our ability to accurately diagnose rare CNS tumor subtypes; however, we discovered limitations that warrant careful consideration. While the system demonstrated robust performance on established and well-characterized subtypes, we also recognize that, with the accumulation of more data and advancements in molecular profiling techniques, new subtypes of CNS tumors are continuously being characterized, such as high-grade gliomas with pleomorphic and pseudopapillary features (HPAP) (Pratt et al., 2022). Reliance on existing 450K array-based reference data for training and classification poses a clear limitation for machine-learning models when confronted with data from previously uncharacterized tumor types.

The introduction of the Illumina Infinium MethylationEPIC v2.0 BeadChip triggered an episode of disarray in neuropathology settings due to the discontinuation of specific CpG probes that were crucial for classifying CNS tumor subtypes. We showed that simply omitting non-applicable CpG probes outputs poor prediction performance and generates discordant classifications from previous models. Model drift, a phenomenon where machine learning models become less effective over time due to changes in data distribution, is a potential challenge in the context of evolving technology and data. To mitigate this, diagnostic laboratories utilizing machine-learning tools must implement mechanisms to detect and correct for model drift. By continuously monitoring model performance and integrating new data with reference datasets, it becomes possible to identify shifts in the data landscape and to adjust classification models accordingly. This approach ensures that diagnostic accuracy remains robust and reliable even as new subtypes of diseases may emerge, or existing ones evolve. At our institution, neuro-oncology diagnostic testing is highly integrated, with cases individually discussed among pathologists, molecular geneticists, and clinical team members. Similar multidisciplinary discussion is imperative to monitor model performance and act quickly when there is a depreciation.

## Data Availability

All code and model data has been deployed in a publicly at (https://github.com/alex-markowitz/pipeline-mmaa) and is available as an open-source pipeline via a Docker container (https://hub.docker.com/r/alexmarkowitz/mmaa_pipeline).

## Acknowledgements

We would like to thank Dr. Jaclyn Biegel, Director of the Center for Personalized Medicine at Children’s Hospital Los Angeles, for her helpful discussions of the study design and implementation of our efforts.

## Funding

Authors declare no funding relevant to this manuscript.

## Notes

**Conflicts of interest:** The authors declare no financial or non-financial conflicts of interest.

### Competing Interest Statement

The authors have declared no competing interest.

### Funding Statement

This study did not receive any funding.

### Author Declarations

All data was collected and anonymized in compliance of policies set by Children's Hospital Los Angeles institutional review board.

